# Postpartum Depression During the COVID-19 Pandemic and Associated Factors: A Cross-Sectional Study

**DOI:** 10.1101/2024.09.22.24314126

**Authors:** Mariana Mie Teruya, Gabriel Sant’Ana Carrijo, Gleise Aparecida Moraes Costa, Mariliza Henrique Da Silva, Júlia Ribeiro Targa de Lima, Julia Fontanezzi Sacramento Veltri Costa, Rodolfo Strufaldi, Cristina Ortiz Sobrinho Valete

## Abstract

**Objective:** To study the prevalence of maternal depressive symptoms in the immediate postpartum period (≤ 7 days postpartum) during the COVID-19 pandemic and to identify associated factors.

**Methods:** We performed a cross-sectional analytical study. The study population consisted of postpartum women >18 years old enrolled at the Municipal University Hospital of São Bernardo do Campo. The Edinburgh Postnatal Depression Scale and a questionnaire on sociodemographic characteristics and clinical history were administered to 90 patients in the first week of the immediate postpartum period between June and December 2021.

**Results:** An Edinburgh Postnatal Depression Scale score > 10 was observed in 39 (43.3%) of the postpartum women. From a multivariate analysis, the factors associated with a score > 10 on the scale were: “having no belief/spirituality” with 83% (95% CI 1.20-2.81, p<0.006), “having had more than one pregnancy” with 50% (95% CI 0.29-0.89, p<0.02), “having a previous episode of depression” with 64% (95% CI 1.02-2.65, p<0.04), and “having a previous episode of anxiety” with 83% (95% CI 1.06-3.16, p<0.03). The other factors studied did not have statistical significance.

**Conclusion:** With the COVID-19 pandemic, an increase in the prevalence of depressive symptoms in the immediate postpartum period was observed. The postpartum women who scored > 10 on the EPDS had the following common associated factors: “having no belief/spirituality”, “having had more than one pregnancy”, “having a previous episode of depression”, and “having a previous episode of anxiety”. These findings indicate the need for greater attention from health professionals to these positive factors.

## Introduction

Postpartum depression (PPD), also known as peripartum depression, is a prevalent and debilitating condition affecting a significant number of women after childbirth. It is defined by the presence of major depression criteria associated with pregnancy or up to 4 weeks postpartum, according to the “Diagnostic and Statistical Manual of Mental Disorders (DSM-V) - 2013.” (1). In Brazil, approximately 15% to 28% of women develop PPD (2). Despite its high prevalence, less than 20% of obstetric teams focus on diagnosing and treating mental disorders during prenatal care, which is a predictor for PPD (2,3).

The pathophysiology of PPD is complex and not fully understood, yet its symptoms can profoundly impact the lives of mothers, their babies, and their families(4). In mothers, untreated PPD is associated with weight problems, impaired social relationships, decreased maternal attachment, use of alcohol and/or illicit drugs, persistent depression, and even suicide (4,5).

The COVID-19 pandemic has led to several mandatory prevention regulations, changes in healthcare services, unprecedented unemployment rates, financial stress, and emotional worries. In this context, the global prevalence of PPD ranged from 6.4% to 56.9% (6), a much higher percentage when compared to the pre-pandemic period. In Brazil, about 38.8% of postpartum women had symptoms consistent with PPD, with the risk of PPD increasing by 2.3 times when the postpartum woman was hospitalized due to COVID-19 infection (7). Given the importance of this issue, this study aims to analyze the prevalence of PPD symptoms in the immediate postpartum period and identify associated factors during the COVID-19 pandemic in a Brazilian obstetric health center.

## Methods

This is a cross-sectional study conducted in a hospital located in São Paulo, Brazil, from June 2021 to December 2021. This study was approved by the Faculty of Medicine of ABC ethics committee (CAAE: 43879521.0.0000.0082) and informed consent was obtained from all participants. This health care center is a tertiary teaching hospital accredited by Baby Friendly Hospital Initiative, located in the ABC region. The maternity ward is a reference for high-risk pregnancies. The neonatal intensive care unit has 20 beds, with a multidisciplinary team working in the areas of audiology and speech therapy, occupational therapy, psychology, social work, and physiotherapy, in addition to the medical, nursing, and specialist teams.

Inclusion criteria were women in the first week of postpartum, aged >18 years. Exclusion criteria were patients who did not adequately answer the Edinburgh Postnatal Depression Scale (EPDS) or who did not consent to partaking in the research.

Sociodemographic and clinical characteristics were investigated. The EPDS was applied to all participants, as a screening tool for PPD. The EPDS is a self-applied questionnaire developed by Cox et al (8), composed by ten items with answers in a four-level Likert scale, translated and validated to Brazil by Santos et al (9). The outcome variable considered was PPD, defined according to a positive screening by EPDS ≥10, as suggested by Figueira et al (10). Other data were collected from electronic charts. Whenever necessary, when the EPDS was ≥10 or when asked by themselves, women were encouraged to follow the institution’s mental health service.

Statistical analysis: All statistical analyses were done using Stata software version 13. Sample power was calculated considering 20% difference, by the Wald statistics and the sample comprising 90 participants, resulting in 0.9854. Normality was evaluated by the Shapiro-Wilk test (p-value < 0.001). Results are presented as medians and interquartile range (IQR), frequency, proportions and 95% confidence intervals (95% CI). Crude prevalence ratios (CPr) were calculated by the Poisson regression (robust estimation and log link function) considering the outcome PPD (yes/no). Variables with a p < 0.20 were tested in a multivariate model to estimate the adjusted prevalence ratios (APr). For all analyses, a p < 0.05 was considered statistically significant. This article writing followed the STROBE form for cross-sectional studies (11).

## Results

The study included 90 participants. The median age was 27.5 years (IQR 23-34), 25 (27.8%) self-identified as white, 54 (60.0%) as brown and 11 (12.2%) as black.

Regarding the educational level, 64 (71.1%) had high school diploma and 11 (12.2%) had university degree.

Twenty-eight (31.1%) were married and 30 (33.3%) were single. Regarding religion, 29 (32.2%) were catholic, 37 (41.1%) were evangelical and 17 (18.9%) reported not having a religion. Sixty-eight (77.3%) reported having a spiritual belief. Those who reported not having a spiritual belief believed their lives’ sense was represented by their infants in 10 (45.4%) and their family in 7 (31.8%). Thirteen (14.4%) had chronic disease and took some medicine during pregnancy. Seventy-five (83.3%) reported having a support network. Thirty-seven (41.1%) reported their family income decreased during the pandemic period. Their working conditions changed in 63 (70.0%), and of these, 38 (60.3%) reported that these changes were negative. Sixty-four (71.1%) reported no alcohol intake habits before and during pregnancy and 22 (24.5%) reported alcohol intake before pregnancy with discontinuation upon discovery. Forty (44.4%) planned pregnancy, thirty-eight (42.2%) reported a previous anxiety episode and 16 (17.8%) reported a previous depression episode.

The median body mass index (BMI) in the beginning of pregnancy was 25.0 (IQR 21.7-30.1), in the end it was 30.1 (IQR 24.9-32.9) and median weight gain was 10 kg (IQR 7-13). Twenty-nine (32.2%) had vaginal delivery and 61 (67.8%) were submitted to cesarean section. Eighty-nine were tested for COVID-19 and of these, 33 (37.0%) were positive.

Considering the newborns’ characteristics, 48 (53.3%) were male sex, with median birth weight 3,157g (IQR 2,800-3,605g), median gestational age 39^0/7^ weeks (IQR 38^0/7^-40^0/7^ weeks), 87 (97.7%) reported the newborn was healthy and 79 (87.7%) reported they were in exclusive breastfeeding at that moment. Median EPDS score was 9 (IQR 5-12) and the detailed responses are in **Table 1**.

**Table 1.** The Edinburgh Postnatal Depression Scale results (n=90)

| Questions | n (%) | Questions | n (%) |
| --- | --- | --- | --- |
| **1. I have been able to laugh and see the funny side of things:**<br>As much as I always could<br>Not quite so much now<br>Definitely not so much now<br>Not at all | 58 (64.4)<br>23 (25.6)<br>8 (8.9)<br>1 (1.1) | **6. Things have been getting to me:**<br>No, I have been coping as well as ever<br>No, most of the time I have coped quite well<br>Yes, sometimes I haven't been coping as well as usual<br>Yes, most of the time I haven't been able to cope at all | 11 (12.2)<br>27 (30.0)<br>36 (40.0)<br>16 (17.8) |
| **2. I have looked forward with enjoyment to things:**<br>As much as ever did<br>Rather less than I used to<br>Definitely less than I used to<br>Hardly at all | 64 (71.1)<br>19 (21.1)<br>6 (6.7)<br>1 (1.1) | **7. I have been so unhappy that I had difficulty sleeping:**<br>Yes, most of the time<br>Yes, sometimes<br>No, not very often<br>No, not at all | 3 (3.3)<br>8 (8.9)<br>35 (38.9)<br>44 (48.9) |
| **3. I have blamed myself unnecessarily when things went wrong:**<br>Yes, most of the time<br>Yes, some of the time<br>Not very often<br>No, never | 15 (16.7)<br>37 (41.1)<br>25 (27.8)<br>13 (14.4) | **8. I have felt sad or miserable:**<br>Yes, most of the time<br>Yes, quite often<br>Not very often<br>No, not at all | 6 (6.7)<br>9 (10.0)<br>34 (37.8)<br>41 (45.5) |
| **4. I have been anxious or worried for no good reason:**<br>No, not at all<br>Hardly ever<br>Yes, sometimes<br>Yes, very often | 17 (18.9)<br>22 (24.4)<br>38 (42.2)<br>13 (14.5) | **9. I have been so unhappy that I have been crying:**<br>Yes, most of the time<br>Yes, quite often<br>Only occasionally<br>No, never | 5 (5.5)<br>6 (6.7)<br>42 (46.7)<br>37 (41.1) |
| **5. I have felt scared or panicky for no good reason:**<br>Yes, quite a lot<br>Yes, sometimes<br>No, not much<br>No, not at all | 6 (6.7)<br>20 (22.2)<br>34 (37.8)<br>30 (33.3) | **10. The thought of harming myself has occurred to me:**<br>Yes, quite often<br>Sometimes<br>Hardly ever<br>Never | 5 (5.6)<br>2 (2.2)<br>6 (6.7)<br>77 (85.5) |

The EPDS ≥ 10 was observed in 39 (43.3%) of participants. Factors associated with this score were “not having a belief”, “previous depression episode” and “previous anxiety episode”. Number of gestations equal to one was associated with lower EPDS (Table 2).

**Table 2.**
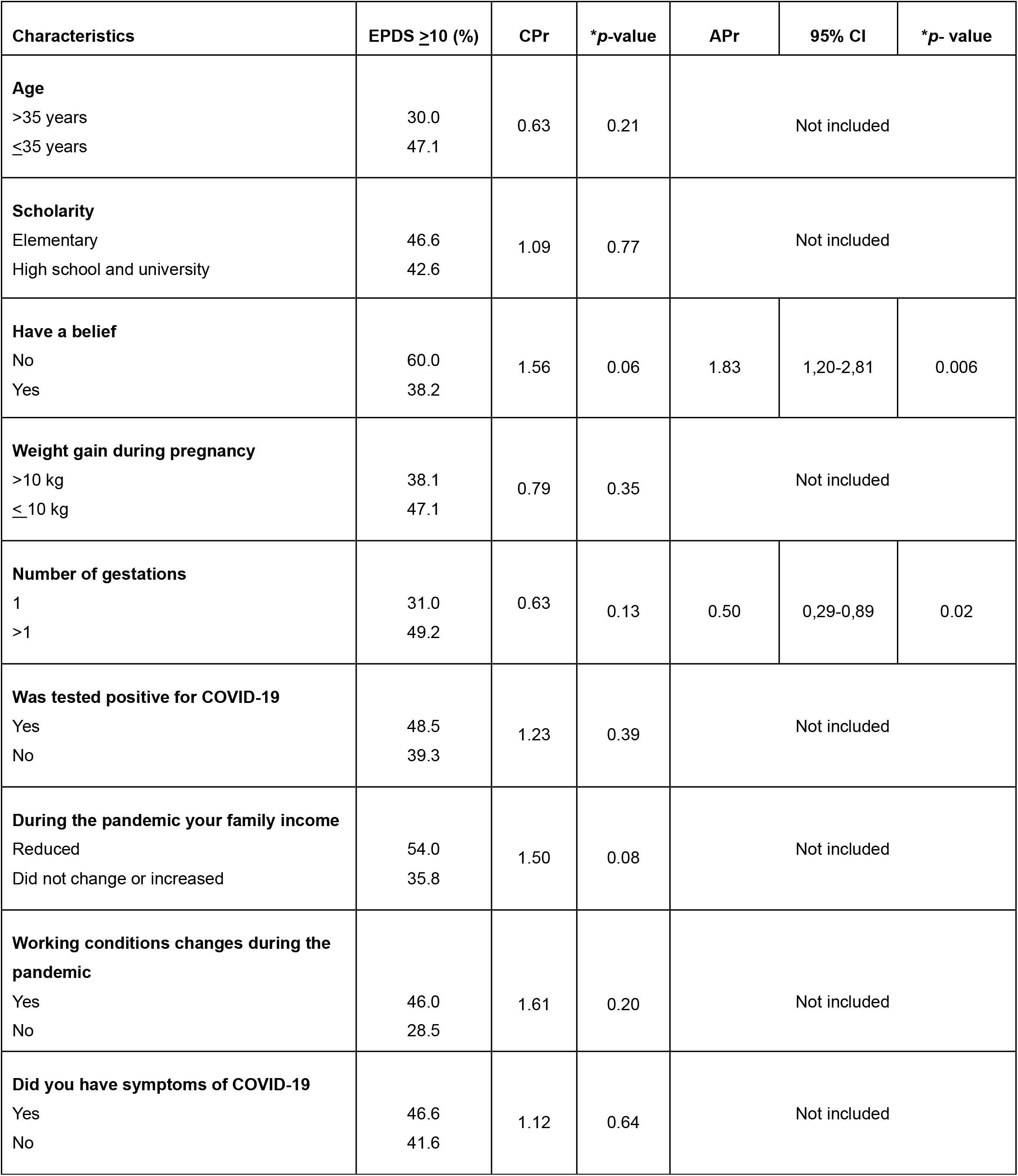
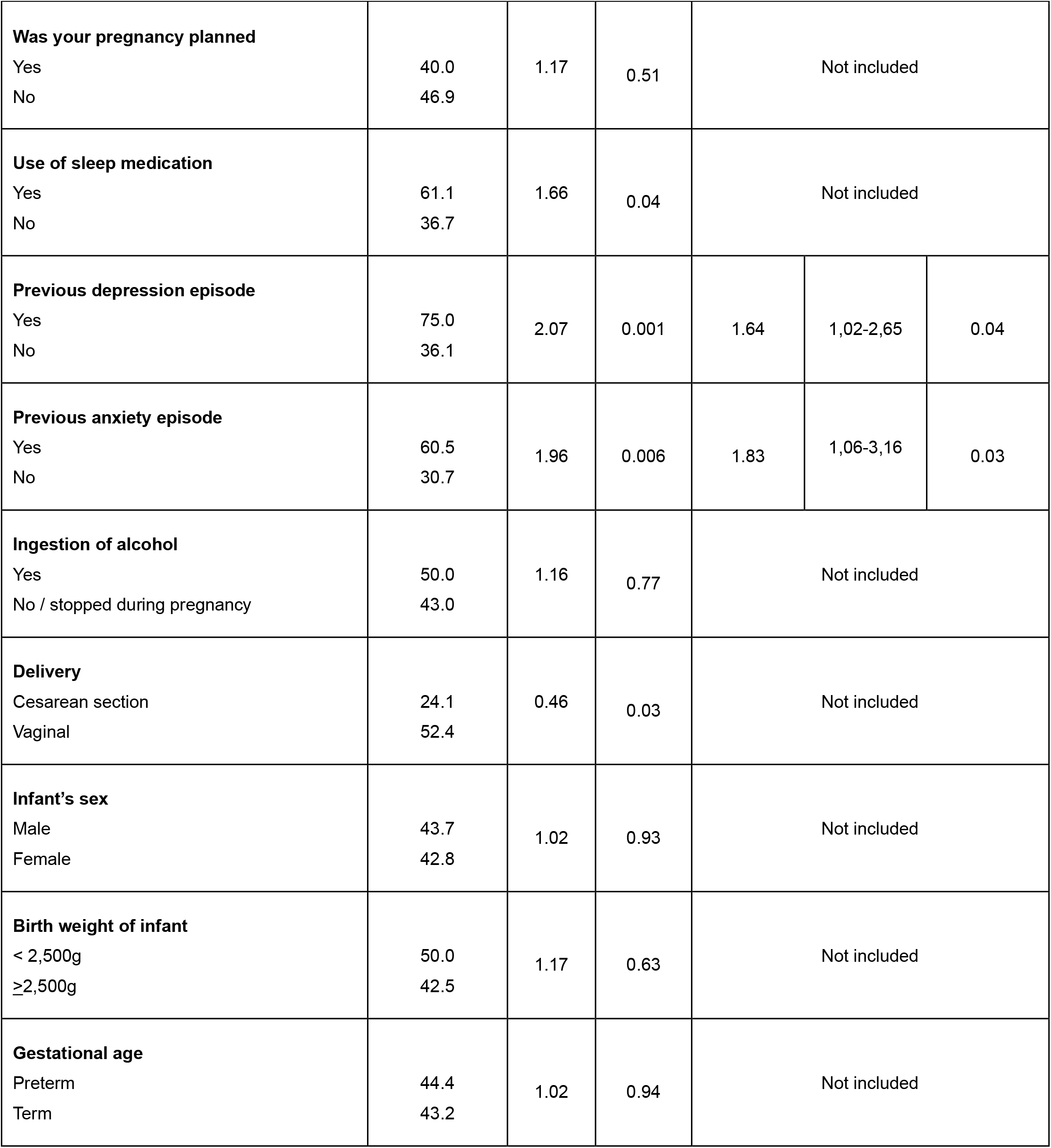
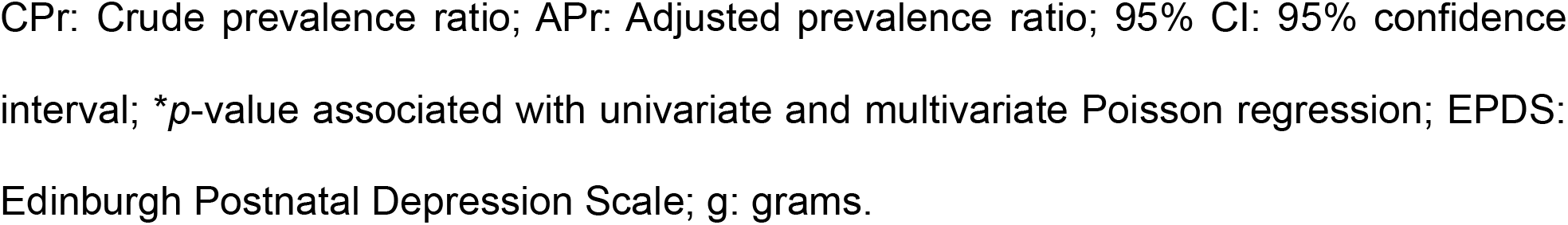
Univariate and multivariate regression model of EDPS > 10 and associated factors (n=90).

## Discussion

In this study, using the EPDS with a cutoff score of ≥10, the prevalence of depressive symptoms in the immediate postpartum period (≤ 7 days postpartum) was found to be 43.3% in the context of the COVID-19 pandemic. Pre-pandemic Brazilian studies that assessed the prevalence of depressive symptoms in the immediate postpartum period reported a prevalence ranging from 6.7% to 24.51% using the same methodology as this study(12,13). The increase in the prevalence of depressive symptoms in Brazil mirrors the global trend. A systematic review and meta-analysis by Chmielewska B. et al. found an increase in EPDS scores when comparing before and after the COVID-19 pandemic, with a pooled mean difference of 0.42 [95% CI 0.02–0.81; three studies, 2330 and 6517 pregnancies](14). Therefore, both in Brazil and worldwide, there was an almost 50% increase in the prevalence of depressive symptoms in the immediate postpartum period following the onset of the COVID-19 pandemic.

Given the statistical analysis, only the variables “lack of spiritual/religious beliefs,” “having had more than one pregnancy,” “previous episode of depression,” and “previous episode of anxiety” were significant risk factors for depressive symptoms in the immediate postpartum period during the COVID-19 pandemic. These factors will be discussed in the following paragraphs.

According to our multivariate regression model, postpartum women without any spiritual beliefs were 83% more likely to have an EPDS score > 10 in the immediate postpartum period during the COVID-19 pandemic. It is known that religious participation and spiritual behaviors are associated with a lower risk of PPD (15,16) and milder depressive symptoms during this period (15).

During the COVID-19 pandemic, the mental health of the general population was impaired. An integrative review by Cordero et al. showed that religious and spiritual beliefs might be associated with better coping, fewer mental health problems (stress, anxiety, depression), and better well-being during the pandemic (17). Additionally, religiosity and spirituality played an important role in alleviating suffering by minimizing the consequences of social isolation (18).

Regarding postpartum women, there is still a lack of studies that relate religiosity/spiritual practices, mental health in postpartum women, and the COVID-19 pandemic, making this study one of the first to relate religion/spirituality to depressive symptoms in the puerperium during the COVID-19 pandemic.

Given this significant result from this study, healthcare professionals should be attentive when attending to and monitoring postpartum women who have experienced the COVID-19 pandemic. Additionally, further research is needed, as the long-term consequences of depressive symptoms in the context of the COVID-19 pandemic on the health of women and their children are still unknown.

In our results, being multiparous was a significant risk factor for developing depressive symptoms during the COVID-19 pandemic, with multiparous women having a 50% higher. This finding aligns with the literature, which indicates that multiparous women are more vulnerable to PPD compared to their first pregnancy. Pre-pandemic studies already showed that being multiparous increased the likelihood of developing PPD (19– 21). According to a systematic review and meta-analysis conducted during the pandemic (22), this association persisted in the new context.

In Brazilian studies, research conducted in Rio Grande do Sul by Hartmann et al. applied the EPDS to a sample of 2687 postpartum women and concluded that parity of two or more was a risk factor for PPD (20). The relationship between the number of children and PPD may be related to stress and family overload when the woman already has other children (20).

Given these results, healthcare professionals and social workers should be particularly attentive to multiparous women, considering not only perinatal care but also the broader social and familial context in which these women live.

The presence of past episodes of depression was shown to increase the likelihood of developing depressive symptoms in the immediate postpartum period (EPDS ≥10) by 64%. Patients with a history of anxiety had an even higher correlation, with 83% of chance. Thus, past episodes of depression and anxiety were significant risk factors for developing depressive symptoms. This is because patients who have experienced depression in the past share common risk factors with PPD, such as socioeconomic fragility, family or marital problems, and genetic risk factors for depression (23). According to the biopsychosocial model of depression (24), patients with these risk factors are more likely to experience depression when faced with a stressful event, such as the arrival of a new child during the pandemic (25). During the COVID-19 pandemic, it was observed that women reported “the thought of not having my partner with me during birth,” “the thought of my partner having to leave the hospital soon after birth,” and “the thought of being separated from my baby after delivery” as the most frequent anxiety inducing thoughts(26). Considering these findings, we suggest that past episodes of depression and/or anxiety be inquired about during prenatal care. Patients with these risk factors require special attention in the postpartum period

The elevated incidence of PPD among women who gave birth during the pandemic carries significant long-term implications for both maternal and child health. PPD has been strongly linked to reduced exclusive breastfeeding time (27), less engagement in pediatric care visits, and greater negligence in completing the child’s vaccination schedule (4). Furthermore, there is a noted delay in the child’s development up to 18 months of age, as well as a 4.7% higher risk of developing depression in adolescence compared to children of mothers free of PPD symptoms (3). These findings highlight the need for addressing potential complications in children born during the pandemic.

This study has limitations. As a single-center study, it reflects a local perspective and has limited external validity. Also, the cross-sectional design precludes any conclusions about the causality or temporal relationships between the identified risk factors and PPD. On the other hand, the strength of this study relies on a scientific contribution to a better understanding of women’s mental health during the immediate postpartum period during the COVID-19 pandemic.

## CONCLUSIONS

Positive screening for depressive symptoms in the immediate postpartum period was frequent and this was associated with previous anxiety, depressive episodes and number of gestations >1. We highlight that spirituality and religion were found to be protective factors for depressive symptoms and this can be approached by the multidisciplinary team.

## Data Availability

All data produced in the present study are available upon reasonable request to the authors

## Authors contributions

Teruya MM, Carrijo GS, Costa GAM, Silva MH, Valete COS contributed to the research conception, design, performed data collection, and wrote the manuscript. Valete COS performed data analysis. Lima JRT, Costa JFSV performed data collection. Strufaldi R supervised the study.

